# Screening for SARS-CoV-2 by RT-PCR: saliva or nasopharyngeal swab? Systematic review and meta-analysis

**DOI:** 10.1101/2021.02.10.21251508

**Authors:** Nusaïbah Ibrahimi, Agnès Delaunay-Moisan, Catherine Hill, Gwénaël Le Teuff, Jean-François Rupprecht, Jean-Yves Thuret, Dan Chaltiel, Marie-Claude Potier

**Affiliations:** Unité de Biostatistiques et d’Épidémiologie, Institut Gustave Roussy, 39, rue Camille Desmoulins, 94805 Villejuif Cedex, France; Université Paris-Saclay, CEA, CNRS, Institute for Integrative Biology of the Cell (I2BC), 91198, Gif-sur-Yvette cedex, France; Aix Marseille Univ, CNRS, Centre de Physique Théorique, Turing Center for Living Systems, Marseille, France; Institut du Cerveau (ICM), CNRS UMR 7225 – Inserm U1127, Sorbonne Université, Paris, France

## Abstract

Diagnosis of COVID-19 in symptomatic patients and screening of populations for SARS-CoV-2 infection require access to straightforward, low-cost and high-throughput testing. The recommended nasopharyngeal swab tests are limited by the need of trained professionals and specific consumables and this procedure is poorly accepted as a screening method. The use of alternative validated samples such as saliva is thus much awaited.

In order to compare saliva and nasopharyngeal/oropharyngeal samples for the detection of SARS-CoV-2, we designed a meta-analysis searching in PubMed up to December 29th, 2020 with the key words “((SARS-CoV-2 OR COVID-19) AND (saliva OR oral fluid)) NOT (review[Publication Type]” applying the following criteria: records published in peer reviewed scientific journals, in English, with at least 15 nasopharyngeal/orapharyngeal swabs and saliva paired samples tested by RT-PCR, studies with available raw data including numbers of positive and negative tests with the two sampling methods. For all studies, concordance and sensitivity were calculated and then pooled in a random-effects model.

A total of 318 studies were retrieved, of which 49 were eligible, reporting on 16,272 pairs of nasopharyngeal/oropharyngeal and saliva samples. Meta-analysis showed high concordance, 92.6% (95%CI: 89.6-94.8), across studies and pooled sensitivities of 86.7% (95%CI: 83.5-89.3) and 92.2 (95%CI: 89.4-94.4) from saliva and nasopharyngeal/oropharyngeal swabs respectively. Heterogeneity across studies was 80.0% for saliva and 84.0% for nasopharyngeal/oropharyngeal swabs.

Our meta-analysis strongly suggests that saliva could be used for frequent testing of COVID-19 patients and “en masse” screening of populations.

**Author summary:** *Why was this study done?:* Three published meta-analysis comparing SARS-CoV-2 loads in paired saliva and nasopharyngeal samples included only 4, 5 and 16 studies up to December 29th, 2020. We thus searched additional studies in PubMed with the key words “((SARS-CoV-2 OR COVID-19) AND (saliva OR oral fluid)) NOT (review[Publication Type])” applying the following criteria: records published in peer reviewed scientific journals, in English, with at least 15 saliva and nasopharyngeal/orapharyngeal paired samples tested by RT-PCR, studies with available raw data including numbers of positive and negative tests with the two sampling methods.

*What did the researchers do and find?:* Forty-nine published studies were eligible, reporting on 16,272 pairs of saliva and nasopharyngeal/oropharyngeal samples. Our unprecedented meta-analysis showed high concordance (92.6%) across studies and pooled sensitivities of 86.7% and 92.2% from saliva and nasopharyngeal/oropharyngeal swabs respectively.

*What do these findings mean?:* Sensitivity of SARS-CoV-2 RT-PCR detection in saliva samples is above the 80% sensitivity cut-off recommended by health regulatory authorities. Our meta-analysis validates the use of saliva sample for mass screening to combat the COVID-19 pandemic.

## Introduction

Propagation of infections by SARS-CoV-2, the coronavirus causing the COVID-19 pandemic, occurs from asymptomatic as well as symptomatic carriers^1^. To reduce the circulation of the virus in the population, SARS-CoV-2 carriers need to be identified rapidly and isolated as soon as possible, ideally before the onset of symptoms. When the virus has disseminated throughout a whole country, massive testing becomes of utmost urgent importance to combat the pandemic ^2-4^.

The recommended diagnosis of SARS-CoV-2 infection from the World Health Organisation is based on real time RT-qPCR detection of viral RNA in respiratory specimen such as nasopharyngeal swabs (NP), bronchial aspiration (BA), throat swab and sputum^5^ The American Centres For Disease Control and Prevention and the European Centre for Disease Prevention and Control now recommend viral testing from the respiratory system such as nasal or oral swabs or saliva^6,7^. Currently, the French health regulatory authority (*Haute Autorité de Santé*)^8^ only supports saliva samples for the detection of SARS-CoV-2 on symptomatic individuals.

To make the diagnostic acceptable to the greatest number of people, especially asymptomatic, massive testing should be based on a sampling procedure that is inexpensive, easy to set up and well accepted by the population^9^. In contrast to nasopharyngeal swabbing, saliva sampling meets these criteria. Saliva sampling is fast, non-invasive, inexpensive and painless. It does not require trained professional and personal protective equipment nor other material than simple plastic tube, and can be self-administered.

To evaluate saliva sampling for the detection of SARS-CoV-2, we conducted a meta-analysis on studies published in peer-reviewed journals until the 29^th^ of December 2020 comparing the detection of SARS-CoV-2 using RT-PCR on paired nasopharyngeal/oropharyngeal and saliva samples in the same individuals sampled at the same time.

## Methods

### Search Strategy and selection criteria

Literature search in PubMed (https://pubmed-ncbi-nlm-nih-gov) run the 29^th^ of December 2020 with search word ((SARS-CoV-2 OR COVID-19) AND (saliva OR oral fluid)) NOT (review[Publication Type]) identified 315 articles. We included publications if they met the following eligibility criteria:

1. Records published in peer reviewed scientific journals;
2. Records published in English;
3. Data curation based on examination of the titles and abstract, searching for research articles assessing viral RNA presence in saliva vs nasopharyngeal and/or oropharyngeal swabs;
4. Availability of nasopharyngeal (and/or oropharyngeal) swabs and saliva data on specimens sampled on the same individuals at the same time;
5. Detection of SARS-CoV-2 using the same RT-PCR method on both samples;
6. More than 15 individuals included in the study.

## Data analysis

From each eligible article, we extracted: the number of individuals positive for SARS-CoV-2 in both nasopharyngeal/oropharyngeal swabs and saliva (a), those positive only in nasopharyngeal/oropharyngeal swab (b), those positive only in saliva (c) and (d) those negative in both nasopharyngeal swab and saliva (**Table 1**). All articles where we highlighted the extracted data are available on the following link^10^. From these data, we calculated the concordance of the same test (RTqPCR for 48 studies and RTdPCR for 1 study) on the two types of sample (a+d)/(a+b+c+d). We also computed the sensitivity of the test on each type of sample. The estimation of the sensitivity of a test requires a reference diagnosis. Since nasopharyngeal swab sampling has been shown to produce false negatives by RTqPCR^11^, sensitivities for the saliva and the nasopharyngeal swab are defined here respectively as (a+c)/(a+b+c) and (a+b)/(a+b+c), considering as true positive any individual with a positive result on one or the other sample. This definition of a positive individual is also in agreement with the US-CDC and the ECDC directives on SARS-Cov-2 testing,

**Table 1:** Studies comparing SARS-CoV-2 detection in paired saliva and nasopharyngeal samples meeting inclusion criteria.

| Reference | Number of tested individuals |  |  |  |  | Concordance | Reference : S+ ou N+ |  |
| --- | --- | --- | --- | --- | --- | --- | --- | --- |
|  | Saliva +<br>Nasoph. + | Saliva -<br>Nasoph. + | Saliva +<br>Nasoph. - | Saliva -<br>Nasoph. - | Total |  | Sensitivity<br>of S | Sensitivity<br>of N |
|  | a | b | c | d | n=a+b+c+d |  | (a+c)/p | (a+b)/p |
| Aita | 7 | 1 | 0 | 35 | 43 | 97.7% | 87.5% | 100.0% |
| Altawalah | 287 | 57 | 18 | 529 | 891 | 91.6% | 84.3% | 95.0% |
| Azzi | 22 | 4 | 33 | 54 | 113 | 67.3% | 93.2% | 44.1% |
| Babady | 16 | 1 | 1 | 69 | 87 | 97.7% | 94.4% | 94.4% |
| Barat | 30 | 7 | 1 | 421 | 459 | 98.3% | 81.6% | 97.4% |
| Berenger | 52 | 11 | 6 | 6 | 75 | 77.3% | 84.1% | 91.3% |
| Bhattacharya | 53 | 5 | 0 | 16 | 74 | 93.2% | 91.4% | 100.0% |
| Binder | 10 | 1 | 1 | 7 | 19 | 89.5% | 91.7% | 91.7% |
| Borghi | 79 | 28 | 7 | 187 | 301 | 88.4% | 75.4% | 93.9% |
| Byrne | 12 | 2 | 0 | 96 | 110 | 98.2% | 85.7% | 100.0% |
| Cassinari | 8 | 5 | 1 | 17 | 31 | 80.6% | 64.3% | 92.9% |
| Caulley | 34 | 22 | 14 | 1869 | 1939 | 98.1% | 68.6% | 80.0% |
| Chau | 19 | 0 | 1 | 7 | 27 | 96.3% | 100.0% | 95.0% |
| Chen | 49 | 6 | 3 | 0 | 58 | 84.5% | 89.7% | 94.8% |
| Güçlü | 23 | 4 | 4 | 33 | 64 | 87.5% | 87.1% | 87.1% |
| Hanege | 29 | 9 | 0 | 0 | 38 | 76.3% | 76.3% | 100.0% |
| Hanson | 75 | 5 | 6 | 268 | 354 | 96.9% | 94.2% | 93.0% |
| Hasanoglu | 27 | 21 | 3 | 9 | 60 | 60.0% | 58.8% | 94.1% |
| Iwasaki | 8 | 1 | 1 | 66 | 76 | 97.4% | 90.0% | 90.0% |
| Jamal 1 | 44 | 20 | 8 | 19 | 91 | 69.2% | 72.2% | 88.9% |
| Jamal 2 | 42 | 14 | 9 | 10 | 75 | 69.3% | 78.5% | 86.2% |
| Kandel | 39 | 4 | 3 | 383 | 429 | 98.4% | 91.3% | 93.5% |
| Kojima | 20 | 3 | 6 | 16 | 45 | 80.0% | 89.7% | 79.3% |
| Landry | 28 | 5 | 2 | 89 | 124 | 94.4% | 85.7% | 94.3% |
| Leung | 38 | 7 | 13 | 37 | 95 | 78.9% | 87.9% | 77.6% |
| Matic | 15 | 6 | 1 | 52 | 74 | 90.5% | 72.7% | 95.5% |
| McCormick-Baw | 47 | 2 | 1 | 106 | 156 | 98.1% | 96.0% | 98.0% |
| Migueres | 34 | 7 | 3 | 79 | 123 | 91.9% | 84.1% | 93.2% |
| Moreno-Contreras | 19 | 9 | 6 | 37 | 71 | 78.9% | 73.5% | 82.4% |
| Nagura-Ikeda | 84 | 19 | 0 | 0 | 103 | 81.6% | 81.6% | 100.0% |
| Otto | 45 | 0 | 4 | 43 | 92 | 95.7% | 100.0% | 91.8% |
| Pasomsub | 16 | 3 | 2 | 179 | 200 | 97.5% | 85.7% | 90.5% |
| Procop | 38 | 0 | 1 | 177 | 216 | 99.5% | 100.0% | 97.4% |
| Rao | 73 | 11 | 76 | 57 | 217 | 59.9% | 93.1% | 52.5% |
| Sakanashi | 15 | 0 | 4 | 9 | 28 | 85.7% | 100.0% | 78.9% |
| Senok | 19 | 7 | 9 | 366 | 401 | 96.0% | 80.0% | 74.3% |
| Skolimowska | 15 | 3 | 1 | 112 | 131 | 96.9% | 84.2% | 94.7% |
| Sorelle | 32 | 7 | 0 | 44 | 83 | 91.6% | 82.1% | 100.0% |
| Sui | 14 | 0 | 2 | 0 | 16 | 87.5% | 100.0% | 87.5% |
| Torres | 46 | 54 | 8 | 835 | 943 | 93.4% | 50.0% | 92.6% |
| Uwamino | 32 | 15 | 11 | 138 | 196 | 86.7% | 74.1% | 81.0% |
| Vaz | 67 | 4 | 2 | 82 | 155 | 96.1% | 94.5% | 97.3% |
| Vogels | 49 | 5 | 4 | 3776 | 3834 | 99.8% | 91.4% | 93.1% |
| Williams | 33 | 6 | 1 | 49 | 89 | 92.1% | 85.0% | 97.5% |
| Wong | 104 | 18 | 37 | 70 | 229 | 76.0% | 88.7% | 76.7% |
| Wyllie | 34 | 9 | 13 | 13 | 69 | 68.1% | 83.9% | 76.8% |
| Yee | 69 | 18 | 10 | 203 | 300 | 90.7% | 81.4% | 89.7% |
| Yokota | 42 | 4 | 6 | 1872 | 1924 | 99.5% | 92.3% | 88.5% |
| Zhu | 382 | 60 | 15 | 487 | 944 | 92.1% | 86.9% | 96.7% |
| Total | 2375 | 510 | 358 | 13029 | 16272 | 92.6% | 84.3% | 89.0% |

The overall concordance and sensitivities have been estimated in a meta-analysis via Generalized Linear Mixed Models (GLMM)^12,13^, using a fixed-effect model, and also a random-effect model in case of an over dispersion of the observations. Dispersion of effect sizes was evaluated using the Higgins *I*^*2*^ estimate of heterogeneity along with the Cochran’s Q in fixed-effect models^14^ and using the tau^2^ estimate (between-study variance which is the variance of the distribution of true effect size) in random-effects models. The tau^2^ was calculated using the maximum likelihood estimator. As some studies have a sample size too small for the normality hypothesis, confidence intervals for each study were computed using the Clopper-Pearson method^15^ also called “exact” binomial interval.

Those for the overall estimates are based on normal approximation. Results are presented as forest plots.

All analyses were done with R version 4.0.3, using the package “meta” version 4.15-1 (2020-09-30) ^16,17^.

## Results

Forty-six studies comparing SARS-CoV-2 loads in NP swabs and saliva samples collected concurrently in the same individuals using the same technique and providing positivity and negativity in both samples have been identified in PubMed^18-63^. We also included 3 articles not identified in the original Pubmed keywords search while fulfilling eligibility criteria (**Table 1 and Figure 1**)^64-66^. In total 16,272 paired samples were analysed. The number of paired samples per study varied between 16 and 3834. Meta-analysis showed an overall concordance of 92.6% (95%CI: 89.6-94.8) across studies (**Figure 2**).

**Figure 1:**
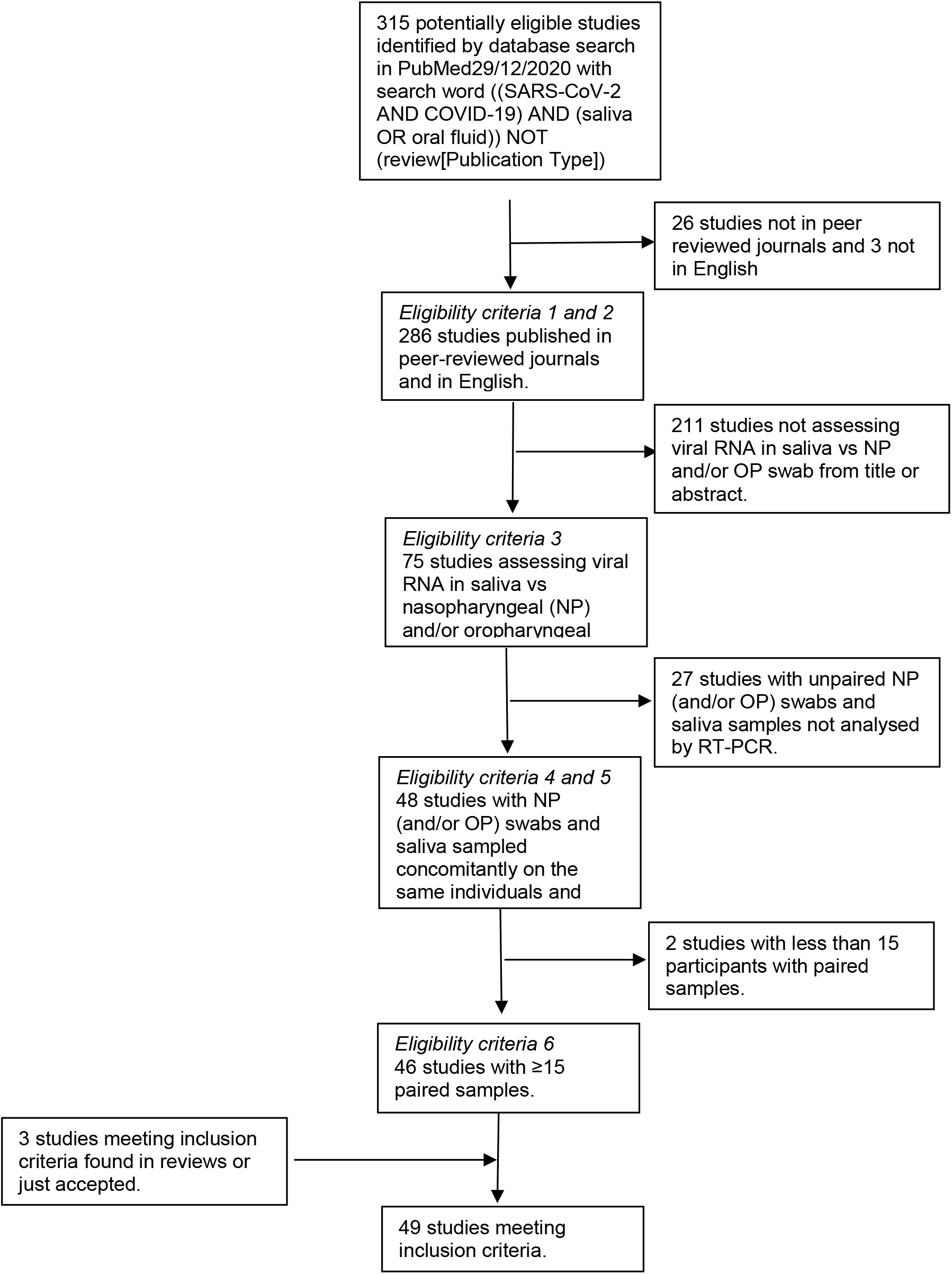
Evidence search and selection.

**Figure 2:**
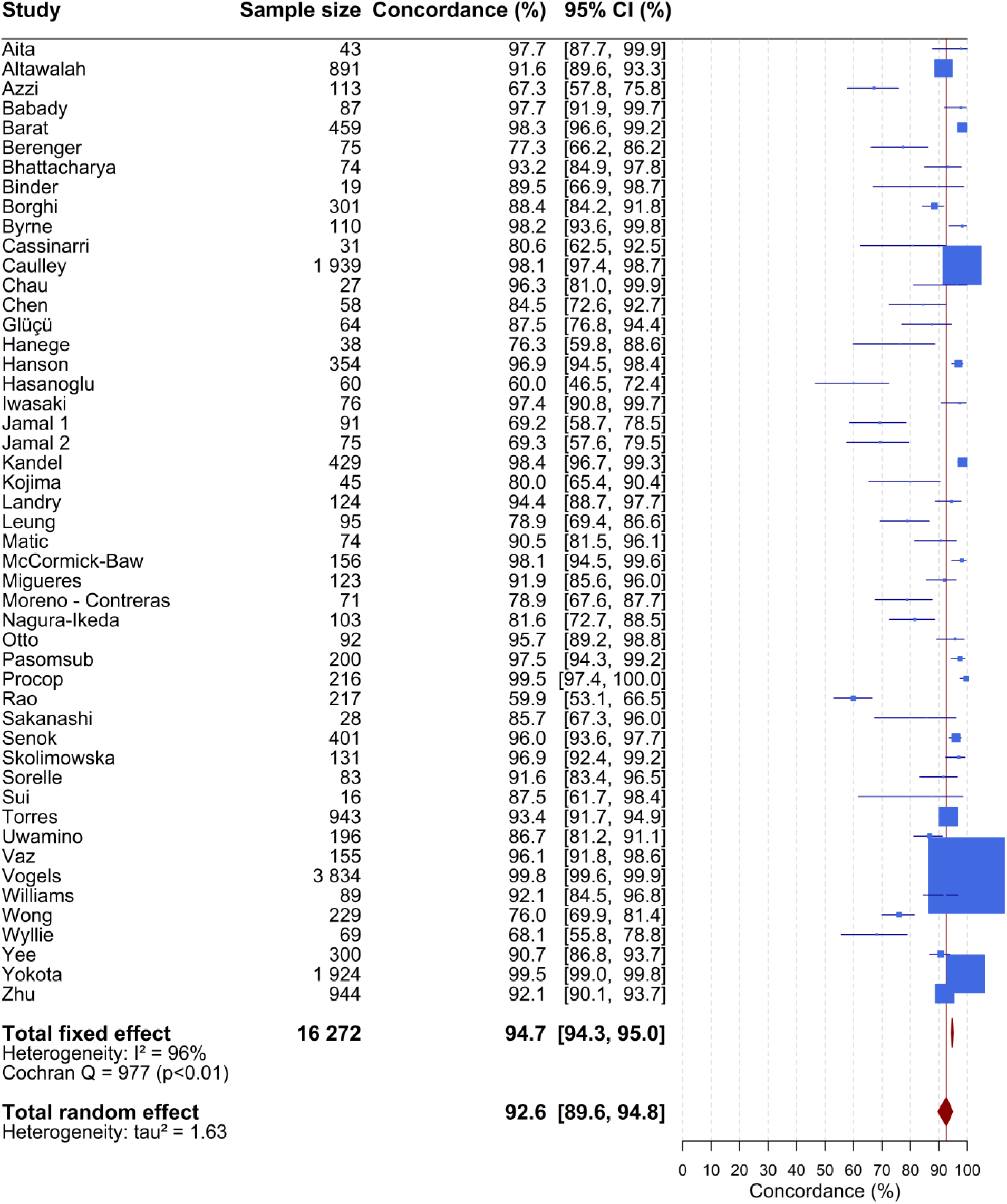
Forest plot of the concordance between the results of RTqPCR tests on nasopharyngeal and saliva samples. The confidence intervals for each study are computed using the Clopper-Pearson method. Those for the overall estimates (fixed-effect or random-effect) are based on normal approximation. The blue box size is proportional to the number of positive tests. The red line corresponds to the value of the overall concordance of the random-effect model. This vertical line enables to locate the studies having an estimate concordance higher than 92.6%.

The overall sensitivity of the RTqPCR test from saliva samples was 86.7% (95%CI: 83.5-89.3) (**Figure 3**) versus 92.2% from nasopharyngeal swabs (95%CI: 89.4-94.8) (**Figure 4**). There was no association between the sensitivities of the saliva (**Figure S1**) or of nasopharyngeal swab (**Figure S2**) estimated in each study and the prevalence of the virus in the same study. If the sensitivity of the saliva was lower in populations of asymptomatic individuals than in population of individuals with symptoms, one would expect to observe a lower sensibility of the saliva in the studies with a low prevalence.

**Figure 3:**
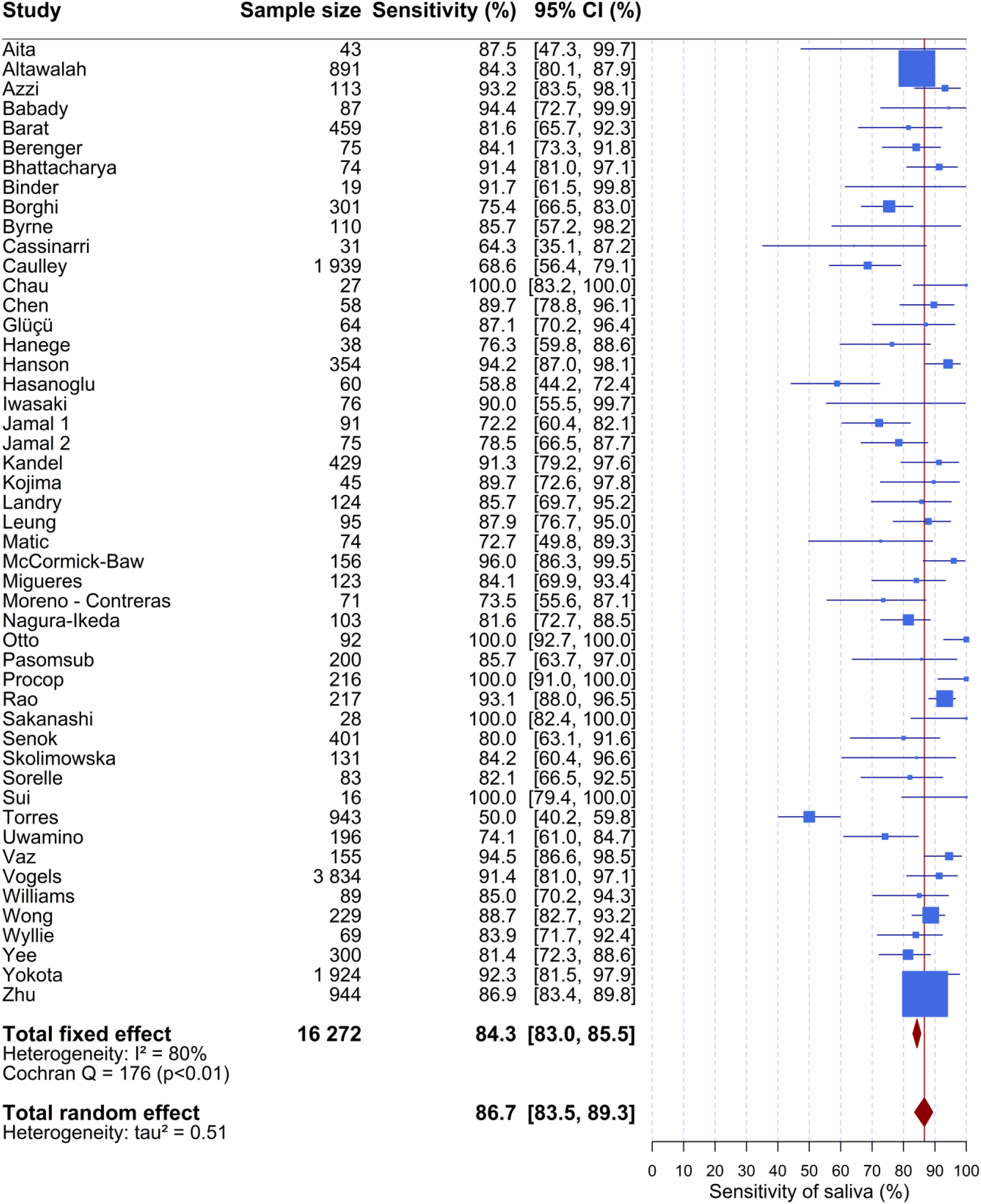
Forest plot of the sensitivity of RTqPCR test on saliva. The confidence intervals for each study are computed using the Clopper-Pearson method. Those for the overall estimates (fixed-effect or random-effect) are based on normal approximation. The blue box size is proportional to the number of positive tests. The difference between fixed-effect or random-effect overall sensitivity (respectively 84.3%, 86.7%) is low. The red line corresponds to the value of the overall sensitivity of the random-effect model. This vertical line enables to locate the studies having an estimate sensitivity higher than 86.7%. The heterogeneity estimator I^2^ is equal to 80%, which means a higher level of heterogeneity.

**Figure 4:**
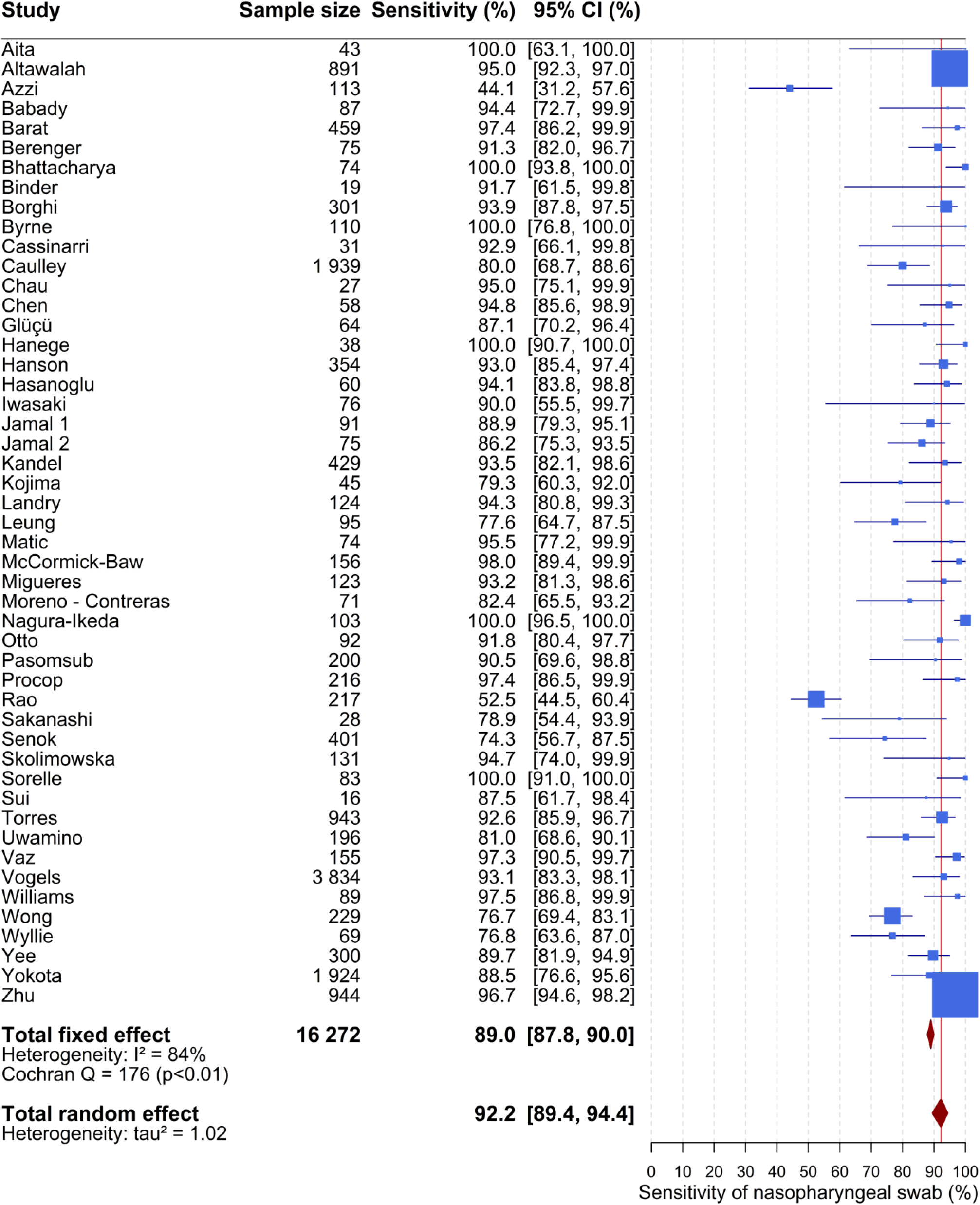
Forest plot of the sensitivity of RTqPCR test on nasopharyngeal sample. The confidence intervals for each study are computed using the Clopper-Pearson method. Those for the overall estimates (fixed-effect or random-effect) are based on the normal approximation. The blue box size is proportional to the number of positive tests. The red line corresponds to the value of the overall sensitivity of the random-effect model. This vertical line enables to locate the studies having an estimate sensitivity higher than 92.2%. The heterogeneity estimator I^2^ is equal to 84%, which means a higher level of heterogeneity.

In our fixed-effect model meta-analysis, saliva gave I^2^ of 80% and nasopharyngeal gave I^2^ of 84%, both corresponding to high heterogeneity. Therefore, a random-effect model was performed to assess the overall sensitivities of the RTqPCR tests, taking into account the fact that the studies do not came from one single population. The variance of saliva was 0.51 and that of nasopharyngeal was 1.02. As a sensitivity analysis, we also used various other methods to estimate the sensitivity, the confidence interval and the tau^2^ and all yielded very similar results.

## Discussion

This meta-analysis reviewed 49 studies and concluded to a high concordance between nasopharyngeal and saliva samples for the detection of SARS-CoV-2 by RT-PCR. Although sensitivity was slightly lower on saliva samples than on nasopharyngeal samples, both values are above the 80% sensitivity cut-off recommended by health regulatory authorities such as the French Haute Autorité de Santé.

For computing concordance and sensitivities, we considered here the reference as SARS-CoV-2 positivity by RT-PCR either in saliva and/or nasopharyngeal samples since the presence of the virus in any sample is indicative of virus carriage.

In the context of mass screening, most participants are asymptomatic. Among the 49 studies analysed, only one included exclusively asymptomatic participants^67^ and 8 studies both symptomatic and asymptomatic participants but it was impossible to separate the data between the two populations^18,21,26,29,51,56,64,66^. One study of contact cases included a larger number of asymptomatic subjects as compared to study of symptomatic subjects^49^. We did not observe any difference in concordance of the tests in these particular studies involving asymptomatic participants (**Table 1**). Formal comparison of nasopharyngeal and saliva samples from asymptomatic individuals is challenging: it would require to screen a large population for a small number of positive cases detected since the prevalence is usually low in this population. On the contrary, the symptomatic population expectedly contains higher percentage of positive subjects, as symptoms usually timely correlates with the highest viral load, which allow an easier comparison of both sampling procedures. Anyhow viral load in saliva of presymptomatic subjects remains in the range of detection of the RTqPCR test for several days both in saliva^68^ and nasopharyngeal samples ^69^. In addition, both asymptomatic and symptomatic subjects appear to be contagious^1^ with similarities in their viral load evolution^70-72^. Moreover, **Figure S3** shows that in France the proportion of positive cases in the symptomatic tested population is consistently about 5 times that of the asymptomatic tested population, independently of any variation of the viral prevalence over time. This and the fact that neither saliva nor nasopharyngeal sensitivities are affected in a screening-like context (the low prevalence being taken as a proxy, **Figure S1 and S2**) strongly predict that viral detection shall exhibit similar performance in both populations.

Our meta-analysis showed large heterogeneity between studies. Sources of heterogeneity are both biological and technical. Biological heterogeneity may come from the fact that a given individual may carry the virus in only one of the saliva or nasopharyngeal specimens, or from the timing of sampling during the course of contamination. Technical heterogeneity comes from differences in the sampling and in RT-PCR methods. Among the 49 studies meeting the inclusion criteria for the meta-analysis, 29 studies report saliva collection in sterile containers (urine tubes or vials) without any additional solution. The other studies diluted the saliva in various viral transport media or phosphate buffer saline with or without bovine serum albumin. Other sources of variability come from differences in the amplified region of SARS-Cov-2 or to different positivity threshold between studies; most studies do not present viral load data but only cycle thresholds (Ct) for specific amplification of SARS-Cov-2 sequences and not even Ct differences (ΔCt) with a human reference gene.

Altogether, our meta-analysis of 49 studies including 16,272 paired samples shows high concordance (92.6%) between nasopharyngeal/oropharyngeal swabs and saliva, with a 5% higher sensitivity for the nasopharyngeal/orapharyngeal (92.2%) as compared to saliva (86.7%). While this might have been a liability in the context of individual diagnosis, this is not such a concern for mass screening, especially given the major advantage of the saliva sampling in terms of logistics. Previous meta-analyses of paired nasopharyngeal and saliva samples included less than 15 peer reviewed studies or preprints and reported average sensitivity of 91%, 85% and 83.4% in 4, 16 and 5 studies respectively ^73-75^. However, these studies used nasopharyngeal positivity as the reference, which did not seem relevant in our context.

To prevent a shortage of analytic reagents and to cut the costs necessarily associated to mass screening strategies, several recent publications have proposed mass testing methods based on saliva sampling either through extraction-free protocols^76- 78^ or through pre-extraction sample pooling^79-81^. In addition, sample pooling has gained a recognized interest for recurrent screening programs from the Centres of Disease Control recommendations^82^, and surveillance protocols implemented in higher education institutions across the world, e.g. the State University of New York (United States)^83^, Liège University (Belgium)^84^, Heidelberg University (Germany)^85^ as well as at Nottingham University (United Kingdom)^86^.

In conclusion, this meta-analysis conclusively demonstrates that saliva is as valid as nasopharyngeal sampling for the detection of SARS-CoV-2 infections in symptomatic as well as asymptomatic carriers. Contrarily to nasopharyngeal swabs, saliva sampling is simple, fast, non-invasive, inexpensive, painless and it thus uniquely applicable for surveillance, screening and diagnosis.

## Supporting information

PRISMA Checklist

## Data Availability

All analyzed articles have been deposited on a drive

https://drive.google.com/drive/folders/1KkbSYdm9NYDfRIHVgaB4DDIm4gPn6f6w?usp=sharing

## Acknowledgements

We wish to thank the FranceTest group for useful discussions and support: Drs. Philippe Froguel, Eric Karsenti, Franck Molina, Jean Rossier and Claire Wyart. We also thank Yannick Marie and Marc Sanson for setting up of the saliva tests at the ICM.

---

**Appendix Figure S1:**
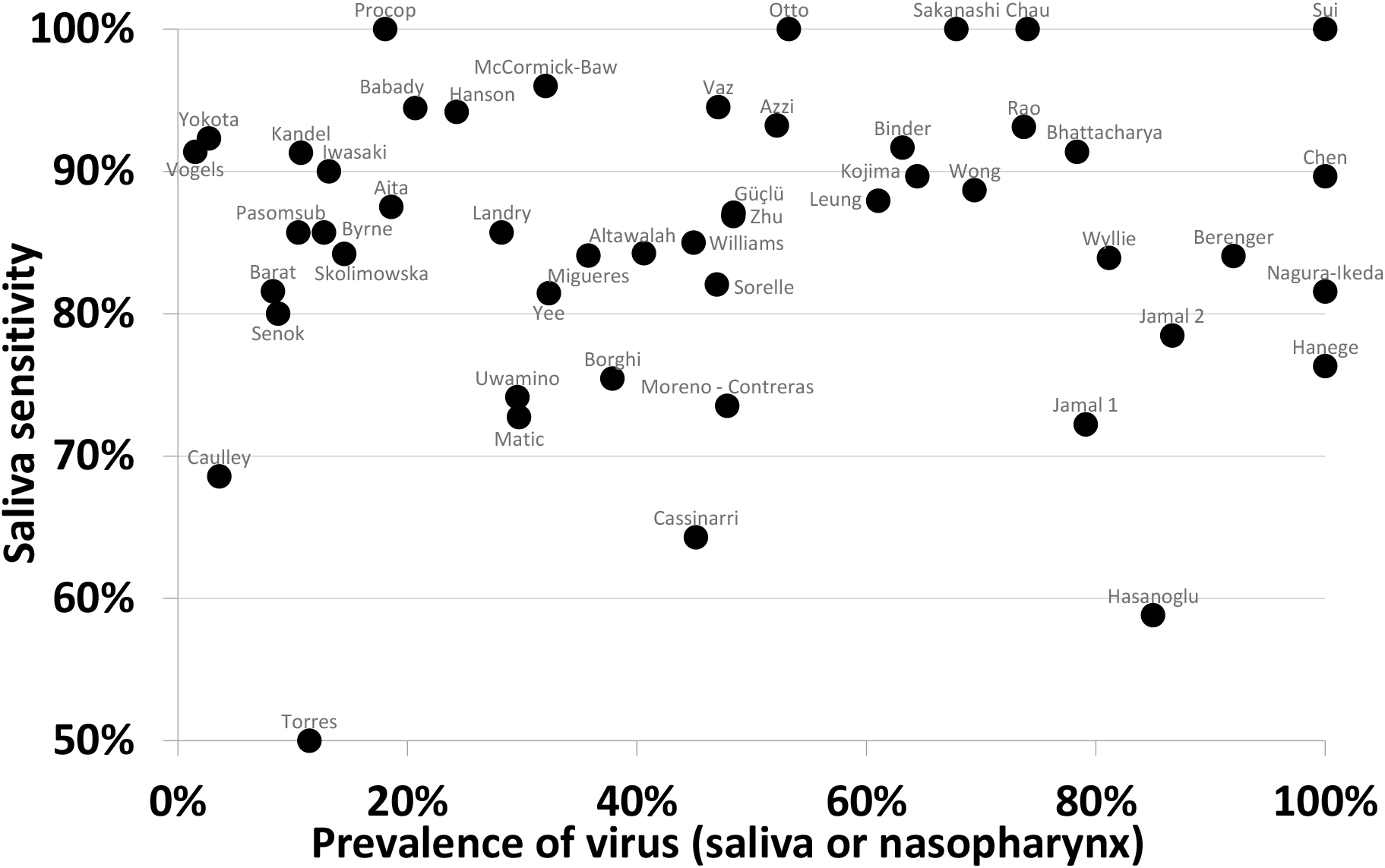
Saliva sensitivity in each study as a function of SARS-CoV2 prevalence.

**Appendix Figure S2:**
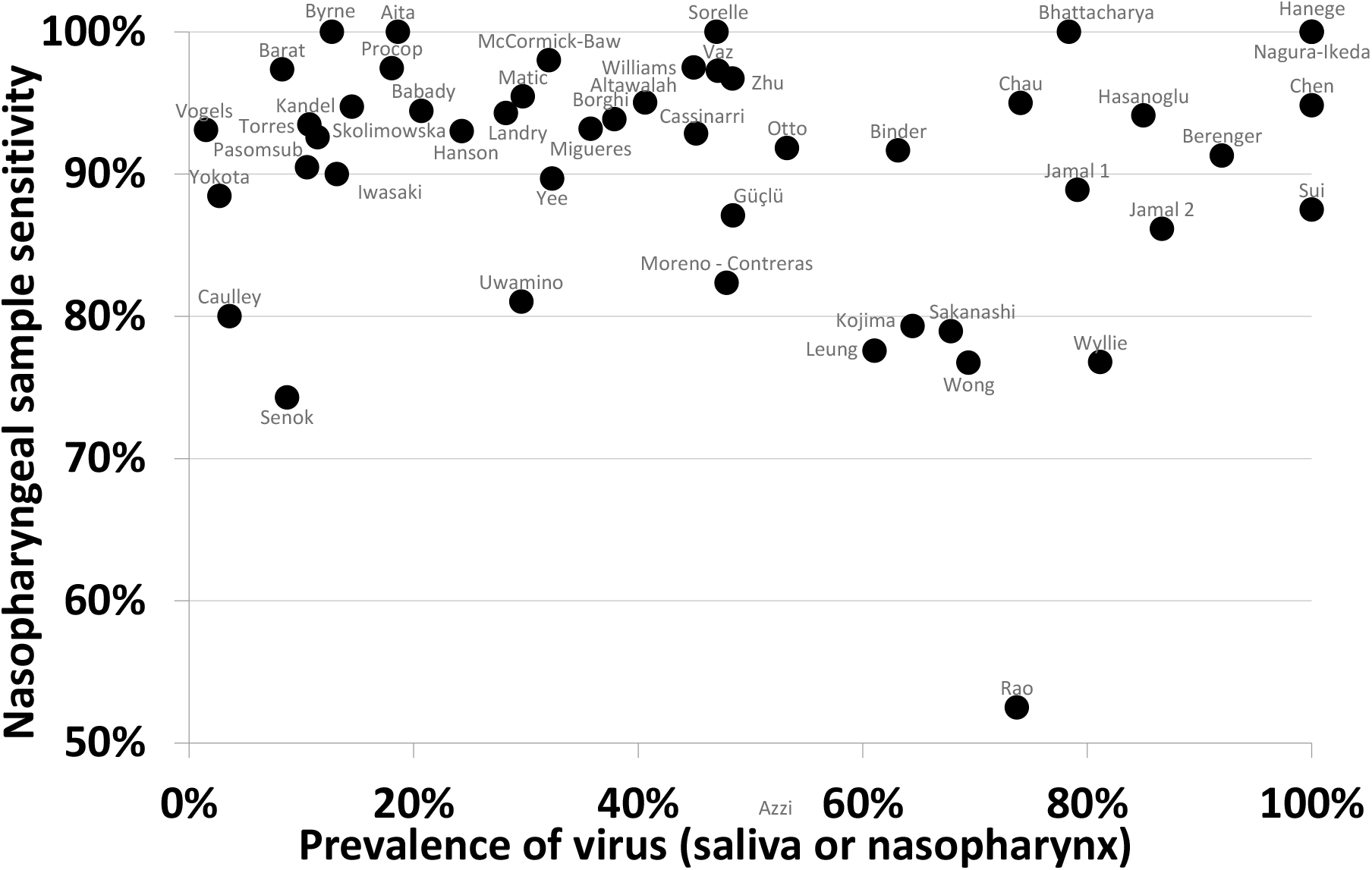
Nasopharyngeal sample sensitivity in each study as a function of SARS-CoV2 prevalence.

**Figure S3:**
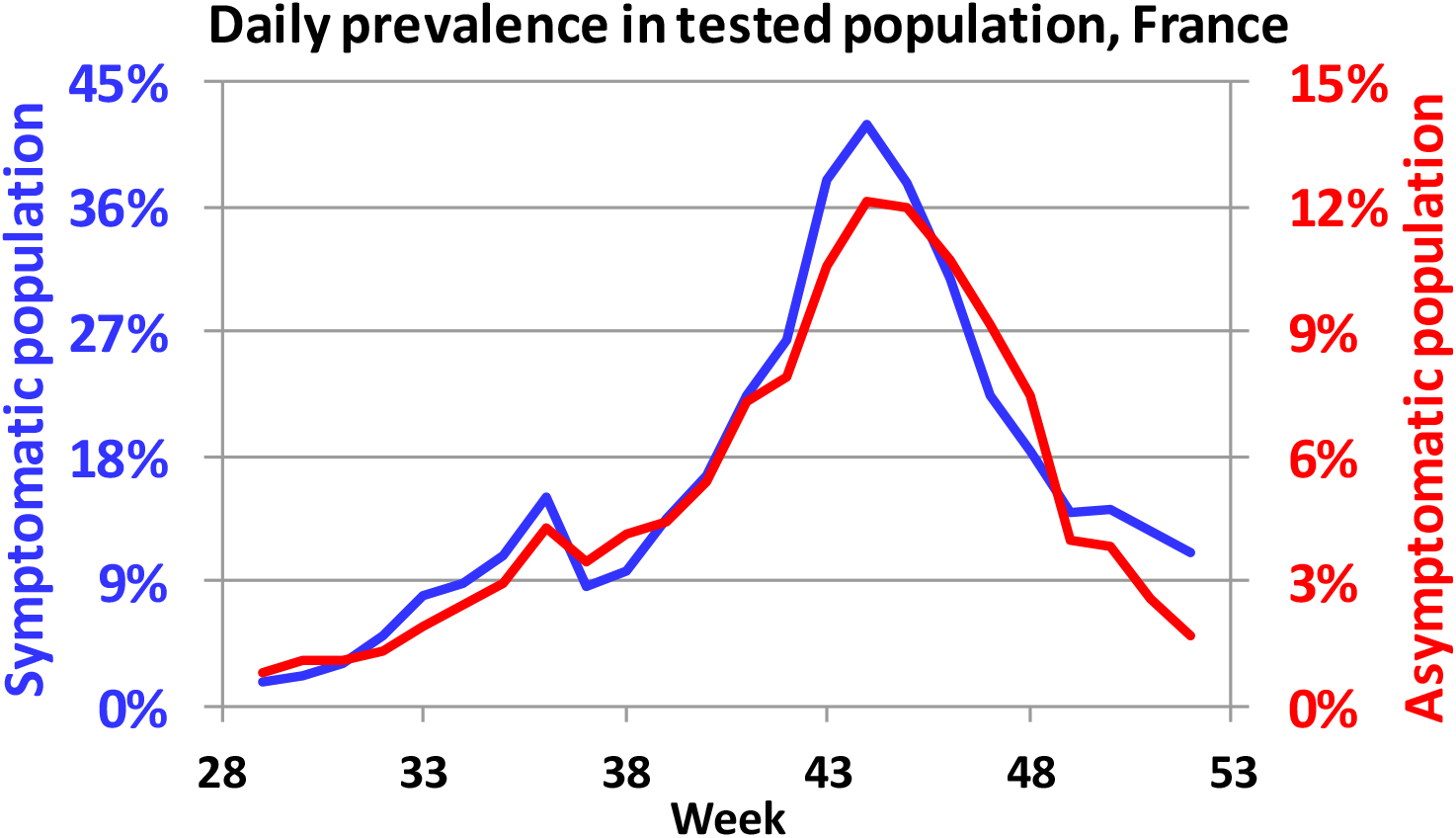
Prevalence in the population tested in France by symptomatic status. Sources: Points épidémiologiques hebdomadaires

